# OvAi Focus: A framework for multiclass segmentation and morphological features extraction in gynecological ultrasound

**DOI:** 10.64898/2026.07.24.26358148

**Authors:** Francesca Salis, Niccolò Tallone, Pio Raffaele Finà, Roberta Massobrio, Rosilari Bellacosa Marotti, Daniele Conti, Luca Fuso, Luca Mariani, Anna Maria Ferrero, Francesca Accomasso, Alessandro Arena, Fulvio Borella, Viola Casula, Stefano Cosma, Tiziana De Grandis, Dan Grisaru, Angelo Lacalandra, Augusto Pereira Sánchez, Renato Seracchioli, Eleonora Robba, Marianna Roccio, Federica Gerace

## Abstract

Ovarian cancer is recognized as the deadliest gynecological malignancy. Diagnosis at advanced stages and the lack of effective screening program lead to poor survival rates, dropping to 17-39 % in stage III-IV diseases. Ultrasound (US) is the primary imaging modality for ovarian structures evaluation, but it is strongly affected by the operator expertise due to the complexity of adnexal masses and the physiological variability of ovarian morphology throughout a woman’s lifecycle. The International Ovarian Tumor Analysis (IOTA) group introduced definitions and predictive tools to standardize gynecological US interpretation. However, these tools still rely on subjective interpretation, thus highlighting the need for more objective solutions. Recent studies have explored artificial intelligence (AI) algorithms for gynecological US, mainly focusing on adnexal masses classification. Conversely, a robust solution supporting the identification and description of healthy and tumoral ovarian structures is still lacking. This paper proposes OvAi Focus, a framework including (i) a segmentation module for the identification of healthy ovaries, plus solid and cystic components of adnexal masses; (ii) a morphology module for the extraction of IOTA-based keywords to describe adnexal masses morphology. Segmentation module results were compared to ground truth masks, showing DICE scores from 0.62 for functional ovary to 0.87 for the whole adnexal mass. Morphology module was tested through interobserver agreement analysis, obtaining Fleiss’ Kappa from 0.16 to 0.57 and Percent Agreement from 47 to 90 %, in line with existing literature. OvAi Focus represents an innovative solution which could help overcoming subjectivity in gynecological US imaging interpretation.

## 1. Introduction

Ovarian cancer is the deadliest gynecological malignancy, characterized by a high mortality rate due to the diagnosis at advanced stages and the absence of effective population-wide screening programs [1, 2]. In 2022, 324 603 new cases and 206 956 deaths were estimated globally, making ovarian cancer the eighth most common cancer and the eighth leading cause of cancer death among women worldwide [3, 4]. The disease typically affects women in mid-to-late adulthood, with a median age at diagnosis around 63 years, and includes several histological subtypes with varying biological behavior and prognosis [3].

The ovary as a healthy organ shows physiological and morphological variability throughout woman’s lifetime, influenced by hormonal cycles and menopause [5, 6, 7, 8]. This dynamic nature increases the difficulty of differentiating normal ovarian structures from pathological findings on imaging. Adnexal masses are a clinically relevant finding in gynecological practice, with 20 % of women developing at least one pelvic mass in their lifetime [9] and occurrence increasing with age, particularly in postmenopausal women [10].

Despite most adnexal masses being benign, a critical challenge remains the reliable identification of lesions with malignant potential at an early stage, also considering their asymptomatic nature in the large majority of cases [11, 9]. Indeed, the 5-years survival rate for patients diagnosed at stage I (around 20-25 % of cases) exceeds 90 %, but it drops to 17-39 % when the disease is diagnosed at stage III or IV (around 75-80 % of cases) [1, 2].

Ultrasound (US) imaging is the primary modality for the evaluation of adnexal pathology due to its safety, real-time visualization, and cost-effectiveness [12]. It allows detailed inspection of internal pelvic structures, including the morphology and vascularization of ovaries and adnexal masses [12]. However, the diagnostic performance of US is strongly influenced by operator experience and subjective assessment, leading to variability in diagnostic consistency and clinical-decision making [13, 14].

To address these challenges, standardized terminology and predictive models have been developed. The International Ovarian Tumor Analysis (IOTA) group introduced a consensus on terms, definitions, and measurements for describing adnexal masses [15] and proposed several predictive tools, including the IOTA Simple Rules [16, 17], the IOTA Simple Rules Risk Assessment [18] and the ADNEX (Assessment of Different NEoplasias in the adneXa) model [19]. In addition, standardized risk stratification systems such as O-RADS (Ovarian-Adnexal Reporting and Data System) were proposed to further unify reporting and management recommendations based on US characteristics [20]. Despite these advances, the application of IOTA terminology and models remains affected by the experience level, posing a concrete challenge for its widespread usage outside specialized centers [21, 22, 23, 24, 25, 26, 27]. In parallel, computational methods have been explored to enhance objectivity and reproducibility. Logistic regression models based on IOTA terms and definitions [28, 29] and radiomics approaches extracting high-dimensional US features have shown promising results in distinguishing benign from malignant lesions [30]. However, they still require manual delineation or feature extraction by experts, thus limiting the clinical applicability of those solutions.

In recent years, artificial intelligence (AI) has emerged as a revolutionary tool in medical image analysis [31]. Several studies focused the attention on the classification of adnexal masses as benign or malignant based on US images, sometimes including segmentation as a preliminary step of the pipeline [32, 33, 34, 35, 36, 37]. Nevertheless, performance evaluation is often limited to the final classification outcome without fully assessing the segmentation quality, which is a crucial step for subsequent feature extraction and interpretation.

Despite the latest scientific works on AI-based segmentation approaches using different imaging techniques [38, 39, 40, 41, 42, 43], existing works based on US are mainly related to the delineation of overall lesion boundaries [36, 44, 37, 45, 46]. The separation of their internal cystic and solid components has not been explored at all, nor fully investigated and validated [45]. A similar trend can be observed for ovaries segmentation: while different studies address follicles segmentation to monitor fertility and potential abnormalities [47, 48, 49, 50], the identification of the whole healthy ovary area remains underexplored [37]. For what concerns IOTA terms and definitions, despite their documented clinical value in describing adnexal masses [51, 52, 53, 54, 55, 56], a valuable solution for their automatic extraction is still missing. Recently, Geysels and colleagues [57] proposed ADNEX-AI, a deep-learning system that automatically segments four ADNEX predictors (lesion, papillary projections, locules and solid tissue) used as input for ADNEX model to predict the malignancy level of adnexal masses. The work illustrates ADNEX-AI segmentation performance for the 5-fold cross-validation dataset, but a proper validation on the test set is lacking, thus preventing a robust generalization capability assessment. Indeed, cross-validation provides an optimistic biased estimation of model’s generalizability, which should be comfirmed on an external test set. Moreover, the algorithm was trained and tested on images acquired using US machines from a single manufacturer (GE Healthcare) and manually selected by an expert operator, which may limit the external validity of the findings.

Notably, the current literature is primarily verticalized on adnexal masses classification, without addressing the need for accurate and objective segmentation and description of both healthy and pathological structures, which is a mandatory prerequisite for the correct patient management. To the best of our knowledge, a validated method based on multi-class segmentation of US images that simultaneously separates solid and cystic components within the mass and segments the ovary is lacking. Furthermore, no established method has been proposed for the automated extraction of US features directly aligned with IOTA definitions, which could help in overcoming the existing limitations.

To address these gaps, the present study proposes a novel AI-based software medical device for gynecological US named OvAi Focus (OvAi Focus, SynDiag s.r.l., Italy) that (i) applies a denoising algorithm to reduce Doppler artifacts [58], (ii) distinguishes solid and cystic components of adnexal masses, (iii) segments the functional ovary (including both healthy ovaries and ovaries with a functional cyst), and (iv) automatically extracts four clinically relevant keywords based on IOTA terms and definitions to describe adnexal masses. Each algorithm was rigorously validated on the test set, demonstrating both technical robustness and potential clinical utility. The rest of the paper is structured as follows: Section 2 reviews the state of the art, while Section 3 introduces the OvAi Focus medical device and describes the Segmentation and Morphology modules from an algorithmic perspective. Section 4 presents the experimental design, including the dataset used for model training and testing, the evaluation strategy, and the data analysis procedures. Section 5 reports the quantitative results, which are discussed in Section 6 in comparison with the existing literature. Finally, Section 7 concludes the paper and outlines future research directions.

## 2. Related Work

### 2.1. Segmentation of adnexal masses and ovaries

To date, only a limited number of studies have addressed the automatic segmentation of ovaries and adnexal masses in US images using deep learning techniques, with most approaches focusing on the delineation of the lesion as a whole rather than on its internal component [59, 60, 61, 62]. Barcroft et al. [36] proposed an end-to-end machine learning framework combining deep learning-based segmentation with radiomics features for the classification of adnexal masses. Their work primarily targets the delineation of the entire mass, with the best segmentation model achieving a Dice coefficient (DICE) in the range of 0.85 - 0.88 across development, validation, and external test sets. While the results demonstrate good generalization capabilities, the segmentation is limited to the outer boundaries of the lesion, without differentiating solid and cystic components. Whitney and colleagues [45] proposed a U-Net–based model for adnexal mass segmentation, followed by fuzzy c-means clustering aimed at separating hypoechoic (cystic) and hyperechoic (solid) regions within the mass. Although the authors report a median DICE of approximately 0.91 for mass segmentation, quantitative performance metrics for the solid and cystic component segmentation are not provided, thus preventing a thorough assessment of the reliability of the proposed components separation. Dai et al. [37] developed a multicenter deep learning pipeline for adnexal mass diagnosis from US images, including a dedicated segmentation module (OvaMTA-Seg). The segmentation model achieved DICEs of 0.887 on internal validation and 0.819 on external validation datasets for full mass delineation. The authors also reported results for healthy ovary segmentation on static images, with DICEs of 0.69 *±* 0.26 on an internal test set and 0.68 *±* 0.27 on an external test set. However, these results were presented in the supplementary material and were not further discussed in the main body of the paper. Zhao et al. [44] introduced the MMOTU dataset, a large multi-modality adnexal masses US dataset, and proposed a dual-stream semantic segmentation network to mitigate cross-domain variability between B-mode and contrast-enhanced US, proposing several approaches. The best solution, based on a DANet architecture, achieved an Intersection over Union (IoU) of 82.6 %. While this work highlights the importance of domain adaptation for ovarian US segmentation, the proposed method is again limited to whole-mass segmentation.

Overall, existing studies predominantly focus on segmenting adnexal masses as a single structure. Compared with these approaches, the OvAi Focus segmentation module addresses both functional ovaries and adnexal lesions and explicitly separates solid and cystic components, providing a structured basis for downstream IOTA-compliant morphological analysis.

### 2.2. Morphological description of adnexal masses

According to the IOTA terminology, the relative amount of solid tissue represents a key morphological feature for adnexal lesions assessment [15]. In current clinical practice, this evaluation is performed qualitatively by clinicians through visual inspection of US images. To date, automated quantification of the relative solid component and its rule-based categorization remain largely unexplored in the literature. Whitney et al. [45] segmented internal mass components into hypoechoic (cystic) and hyperechoic (solid) regions; however, they neither provided quantitative validation nor rule-based classification based on solid–cystic proportions. More recently, Geysels et al. [57] proposed a deep learning framework for automatic extraction of several ADNEX predictors (lesion, locules, solid tissue, and papillary projections). However, the solid component is estimated according to ADNEX definition (i.e., largest solid-component diameter relative to lesion diameter) rather than the relative amount of solid tissue described in the IOTA terminology, without providing a rule-based lesion categorization. OvAi Focus addresses this limitation by introducing a simple and reproducible approach based on the solid-pixel ratio derived from segmentation outputs, enabling objective morphological categorization within an IOTA-aligned framework.

Acoustic shadows constitute another relevant morphological feature in ovarian tumor assessment and contribute to malignancy risk stratification [55]. Although several algorithmic approaches for shadow detection have been proposed, they have primarily been developed in non-gynecological contexts [63, 64, 65, 66, 67, 68, 69]. Hellier et al. [67] developed an automatic geometric and statistical method by combining fan-beam geometry with intensity-based statistical analysis to detect acoustic shadows in intraoperative brain US. Karamalis et al. [68] introduced US confidence maps based on random walks, providing per-pixel estimates of signal reliability that indirectly capture shadowed regions, as areas affected by attenuation or reflection typically exhibit low confidence values. Park and Kim [69] proposed a simpler image-based approach based on local intensity differences along scanlines, demonstrating effective shadow detection in abdominal imaging. To the best of our knowledge, none of these methods has been specifically designed or validated for gynecological or adnexal US. The OvAi Focus acoustic shadow algorithm builds upon these ideas by integrating geometric scanline analysis with confidence-map-based validation, extending shadow detection to gynecological US and improving robustness to noise and anatomical variability. Within IOTA framework, margin regularity contributes to the morphological characterization of ovarian tumors and supports malignancy risk assessment [15]. Automated assessment of lesion margin regularity has been extensively studied in breast US computer-aided diagnosis [70, 71, 72, 73]. Huang et al. [72] extracted contour-based geometric descriptors from breast lesions and applied machine learning classifiers to distinguish benign from malignant tumors, demonstrating that boundary irregularity is a strong discriminative feature in US imaging. More recently, Nairuz et al. [73] proposed frequency-domain irregularity features derived from lesion boundaries in breast US images, achieving improved classification performance when combined with support vector machines. Despite the clinical relevance of irregular margins in ovarian US, automated margin analysis in this domain remains largely unexplored. The OvAi Focus Morphology module builds on concepts from breast computer-aided diagnosis by extracting geometric and statistical features from segmented lesion contours and classifying margin regularity using a logistic regression model tailored to adnexal lesions.

A further component of IOTA framework is the qualitative lesion classification, which integrates septation and solid components into five descriptive categories (unilocular, unilocular-solid, multilocular, multilocular-solid, and solid) [15]. Despite its clinical relevance, no prior studies have proposed automated methods specifically aimed at extracting this feature from US images. Although ADNEX-AI [57] automatically extracts ultrasound predictors for the ADNEX risk model, including the number of locules and the solid component, it does not explicitly derive the IOTA lesion classification. As discussed above, most existing AI-based approaches primarily focus on global lesion detection or benign–malignant classification. To the best of our knowledge, the OvAi Focus Classification network represents one of the first attempts to automatically derive IOTA-compliant classification from US images using a dedicated Convolutional Neural Network (CNN).

## 3. OvAi Focus

In this section we describe the proposed medical device with a focus on the AI-based framework and its development.

OvAi Focus is a stand-alone software medical device designed to support medical doctors in the anatomical description of functional ovaries (including both healthy ovaries and ovaries including a functional cyst) and in the characterization of adnexal lesions in pre and postmenopausal women. The system includes the following main blocks, outlined in Figure 1:

- A Segmentation module, for the identification and segmentation of functional ovaries and adnexal masses, including the distinction between solid and cystic components within each lesion;
- A Morphology module, for the detection of morphological features within the cystic and solid components of the lesion and morphological information extraction;
- A Doppler denoising module, for the improvement of the perceived Doppler signal quality through artifact suppression.

**Figure 1:**
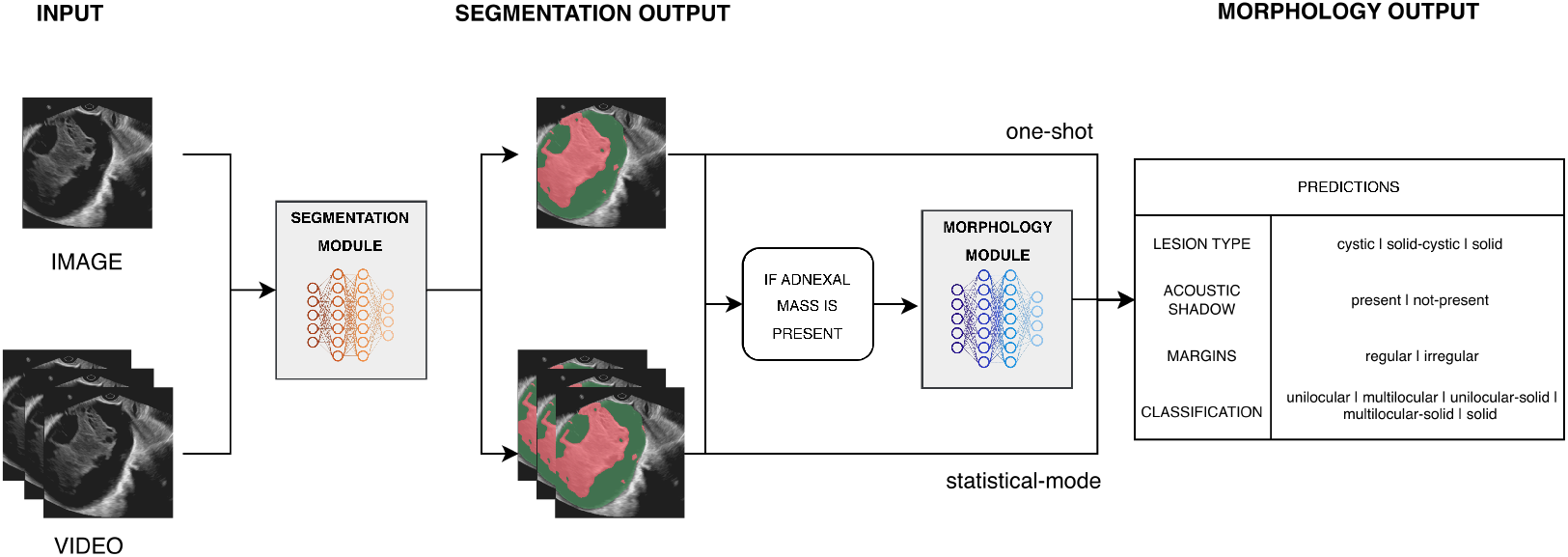
This figure should report the workflow of OvAi Focus AI, with an overview on the required input, the main modules (segmentation and morphology) and the visualized output.

The Doppler functionality has already been described and investigated in a previous study [58], whereas the present work provides a detailed description and analysis of the Segmentation and Morphology functionalities.

The principle of Segmentation module can be summarized as follows:

1. The user provides as input a B-mode US image or video interval in which a functional ovary and/or ovarian lesion may be present. The input data can be provided in DICOM, JPEG, PNG, AVI and MP4 formats;
2. All input items should respect a minimum resolution quality threshold (scale 448×448). Each frame is read and preprocessed individually (e.g., normalization, resizing) to meet quality requirements of the medical device;
3. For each image/frame, the software segments the boundaries of the areas of interest, i.e. functional ovary, cystic areas of adnexal lesion and solid areas of adnexal lesion;
4. For each image/frame, the system highlights:

- In case of functional ovary, the full ovarian structure;
- In case of adnexal mass, the liquid anechoic areas, liquid echoic areas and solid areas associated with the lesion. Liquid anechoic and echoic areas are differentiated using a pixel intensity threshold to improve visualization quality.

The Morphology module is launched after the execution of Segmentation module and works according to the following steps:

1. The output of the segmentation module, i.e. the predicted masks of cystic components of adnexal masses and solid components of adnexal masses, is used as input for the morphology module;
2. For each image/frame, the algorithm assigns a value to four keywords, compliant with IOTA terms and definitions [15], specifically:
  - Lesion type: cystic/solid-cystic/solid;
  - Acoustic shadow: presence/absence;
  - Margins: regular/irregular;
  - Classification: unilocular/multilocular/unilocular-solid/ multilocular-solid/solid.
3. For each image/video, the system summarizes and graphically presents the four IOTA-based keywords.

The output of the two modules is a frame-level set of overlays and measurements that are later aggregated into a video-level summary, if the input is a video, or a single overlay and measure if the input is an image. The system output is generated as on-screen information (numbers, text, images, graphics) and printable reports, which can also be exported as digital documents (e.g., Word, PDF, JPEG).

### 3.1. Segmentation module algorithm

The primary aim of the Segmentation module is to isolate the Region of Interest (ROI) within the US data by predicting pixel-level masks for target structures. Architecturally, this module is designed as a cascade of two distinct sub-modules, illustrated in Figure 2. The first, the Fan-Beam Segmentation module, isolates the active US fan from the image background. The resulting foreground ROI is then processed by the second sub-module, Organ Segmentation, which identifies specific anatomical structures, specifically functional ovaries and adnexal masses, differentiating between solid and non-solid components for the last ones. Detailed descriptions of these modules are provided below.

**Figure 2:**
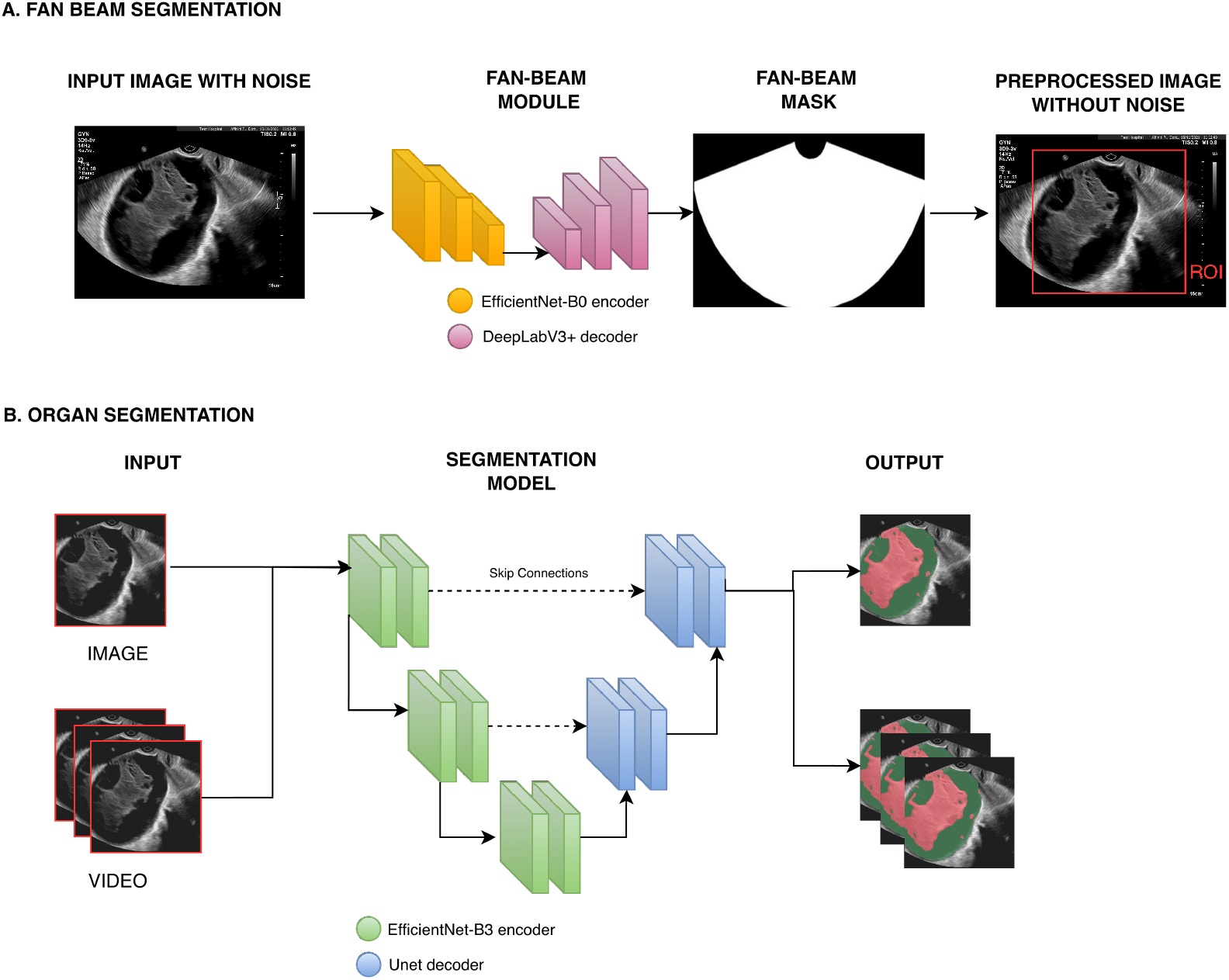
Segmentation algorithm workflow. A. Fan-Beam module; B. Organ Segmentation module.

The Fan-Beam module is built upon the DeepLabV3+ architecture [74], with an EfficientNet-B0 [75] encoder. The network is designed to perform a binary semantic segmentation task, processing raw US imaging data (images or video frames) to distinguish between the foreground (the interior of the fan-beam cone) and the background (the complementary area of the image). Training is conducted in a supervised setting, utilizing pairs of US images and manually annotated ground-truth masks. Further details regarding the training procedure are outlined in Section 3.1.3.

This module is designed to mitigate biases related to Shortcut Learning [76], a known challenge in deep learning. Medical Imaging frequently contains non-anatomical information “burned in” to the pixel data (Figure 2.A1), such as Protected Health Information (PHI) and acquisition settings (e.g., probe orientation markers, depth, and gain values). For US imaging, this information typically resides in the background area outside the fan-beam cone. As demonstrated in prior literature [77, 78], deep learning models may exploit these “non-anatomical cues” to form spurious correlations with the target variable, thereby hindering generalization. To address this, the Fan-Beam module automatically detects (Figure 2.A2) the fan beam cone, allowing the system to discard irrelevant background noise. Consequently, downstream modules, such as organ segmentation and morphology, process only the masked image containing clinically relevant foreground data (Figure 2.A3).

The Organ Segmentation Module is built upon a U-Net-based neural network [79] with an EfficientNet-B3 encoder [75]. This network is trained to perform a multi-class semantic segmentation task, accepting the cropped ROI derived from the fan-beam mask as input (Figure 2.B). Its objective is to segment specific anatomical regions, classifying pixels into one of three categories: healthy ovary, solid adnexal mass components, and cystic adnexal mass components. Like the previous module, this network is trained using a supervised approach with manually annotated ground-truth masks (see Section 3.3 for training details). The output of this sub-module serves two distinct functions. First, the segmented regions are overlaid onto the raw US data to create an augmented video/image displayed for the user. Second, in case of adnexal mass present, the generated masks feed directly into the downstream Morphology Module (Section 3.2). By isolating these structures, the system can extract clinically relevant features as described in the following section.

### 3.2. Morphology module algorithm

The Morphology module takes as input the output of the Segmentation module for clinical cases with at least an adnexal mass, and applies a preliminary filtering step. Frames in which the segmented solid mass covers less than a certain percentage of the total image area are discarded, as they are not considered suitable for morphological analysis. The Morphology module is a suite of two traditional Computer Vision and two Machine Learning algorithms (Figure 3). The working principle of each algorithm is illustrated in the following, while further details regarding the training procedure are outlined in Section 3.3.

**Figure 3:**
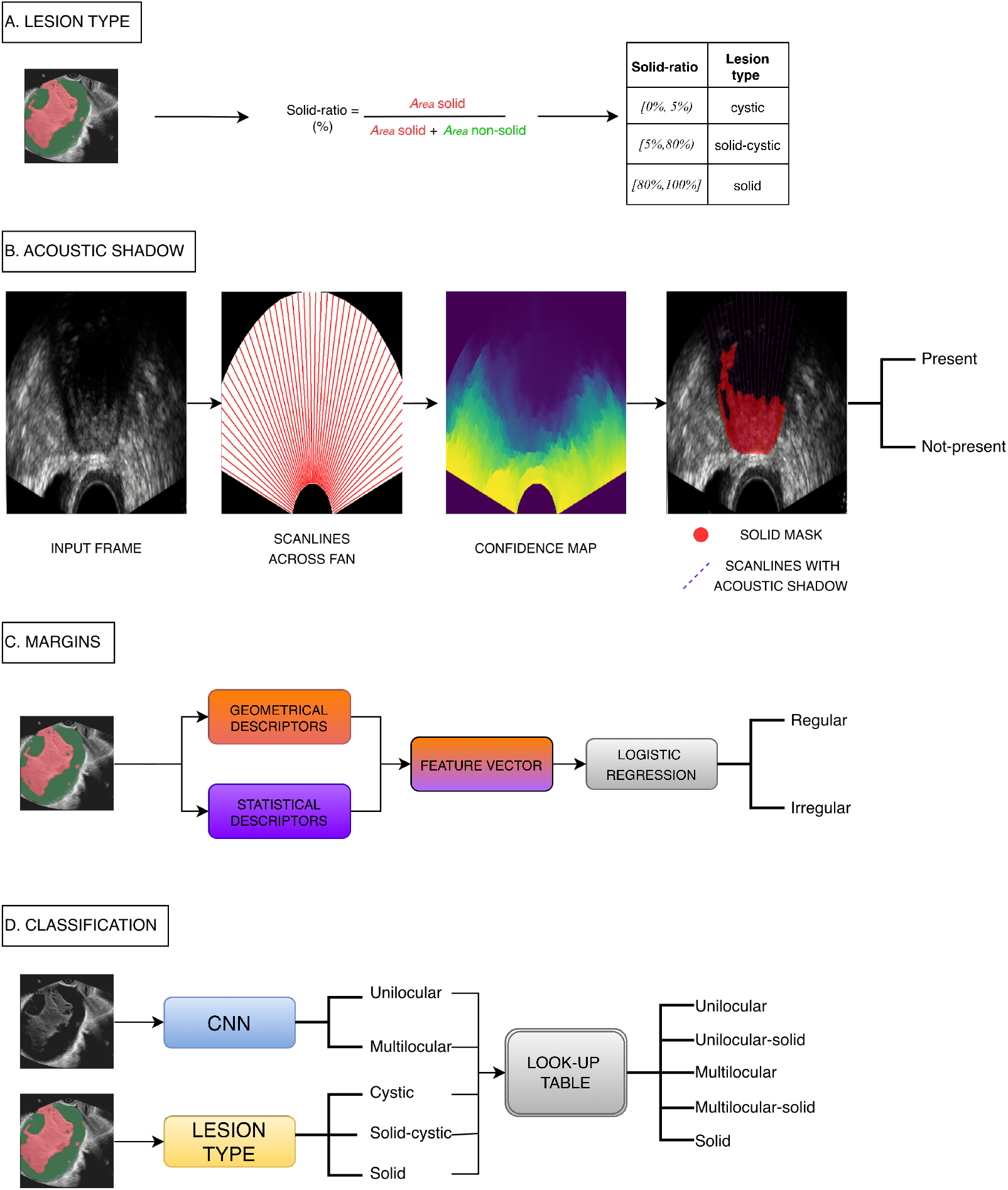
Morphology algorithm workflow. A. Lesion Type; B. Acoustic Shadow; C. Margins; D. Classification

Morphology’s Lesion Type is a classification algorithm designed to characterize ovarian lesions based on their internal composition into three classes: cystic, solid-cystic, or solid (Figure 3.A). This classification is based on the output provided by the OvAi Focus segmentation module for what concerns cystic and solid masks. The method proceeds as follows:

1. For each frame of an input video or image, the so-called solid-ratio, defined as the percentage of the pixels identified by OvAi Focus as solid pixels with respect to the total number of pixels identified by OvAi Focus as part of the ovarian lesion mass, is computed;
2. Each frame is then classified into one of the three lesion types by applying a set of predefined thresholds to the calculated solid-ratio:

- Cystic: solid-ratio < 5 %
- Solid–cystic: 5 % *≤* solid-ratio *<* 80 %
- Solid: solid-ratio *≥* 80 %
3. For video input sequences, the IOTA-based Lesion Type output keyword is determined by computing the statistical mode of the frame-level classifications. For image input, the suggested keyword itself is considered.

Morphology’s Acoustic Shadows is a classification algorithm that uses computer-vision methods to detect clinically relevant acoustic shadowing from US data and the Segmentation module output (Figure 3.B). The method proceeds as follows:

1. The fan-beam geometry of each US image/frame is analyzed to determine transducer orientation relative to the image/frame origin; this step allows extraction of scanlines, defined as radial lines spanning from the near to the far field;
2. A per-pixel US confidence map is generated using the random-walk-based method proposed by Karamalis and colleagues [68], providing a quantitative estimate of US signal transmission reliability within the fan. Low confidence values typically indicate a reduction of signal integrity due to reflection or absorption at solid interfaces, characteristic of acoustic shadowing;
3. For each scanline crossing the solid tissue region identified by the Segmentation module, a confidence profile is obtained by sampling the confidence map along the scanline. A rupture point detection algorithm identifies abrupt drops as candidate acoustic shadow origins. These candidates are then validated by verifying that the subsequent segment of the profile remains consistently low in confidence, ensuring robustness against imaging noise. Validated scanlines are classified as containing an acoustic shadow;
4. Frame-level classification is determined by counting the scanlines with an acoustic shadow. If this count exceeds a predefined threshold, the frame is considered positive for acoustic shadow;
5. For video sequences, the IOTA-based Acoustic Shadow output keyword is obtained as the statistical mode of the frame-level classifications. For image input, the suggested keyword itself is considered.

Morphology’s Margins is a classification algorithm designed to characterize the regularity of ovarian lesion boundaries (Figure 3.C). The core of this algorithm is a logistic regression model trained on geometric and statistical descriptors based on the ovarian lesion regions identified by the Segmentation module, together with ground truth labels. The model proceeds as follows:

1. For each input frame/image, the system extracts two descriptor sets from the lesion contours and regions identified by OvAi Focus:

- geometric descriptors that characterize lesion shape;
- statistical descriptors capturing boundary characteristics.
2. These two descriptor sets are concatenated into a single feature vector representing the lesion’s shape and characteristics for that frame or image;
3. The resulting feature vector is supplied to the logistic regression model, which computes the probability of the feature vector belonging to each possible class, thereby outputting the most probable class assignment;
4. For video input sequences, the IOTA-based Margin output keyword is determined by computing the statistical mode of the frame-level classifications.

Morphology’s Classification is a classification algorithm designed to characterize ovarian lesions into five classes (unilocular, unilocular-solid, multilocular, multilocular-solid, solid) (Figure 3.D). The core of the algorithm is a CNN trained on US data with the corresponding ground-truth labels. The method proceeds as follows:

1. During inference, the input US image is provided to the network, which computes the probability of the image belonging to each possible class and outputs the most probable binary classification (unilocular or multilocular);
2. The network output is then combined with the Lesion Type output keyword to derive the final five-level IOTA Classification output. The mapping between these two inputs is defined in the look-up table reported in Table 1;
3. For video input sequences, the IOTA-based Classification output keyword is obtained by computing the statistical mode of the frame-level classifications. For image input, the suggested keyword itself is considered.

**Table 1:** Mapping between the Classification Network output and the corresponding lesion type output keywords.

| Classification Network Output | Lesion Type Output Keyword |  |  |
| --- | --- | --- | --- |
|  | cystic | solid-cystic | solid |
| unilocular | unilocular | unilocular-solid | solid |
| multilocular | multilocular | multilocular-solid | solid |

### 3.3. Training and validation methodology

All machine learning models presented in Sections 3.1 and 3.2 were trained using a supervised learning approach. The training dataset consisted of raw US data (images and video frames) paired with their corresponding pixel-level ground-truth masks or labels. During the training phase, the model parameters (weights) were iteratively adjusted to minimize a loss function, which quantifies the discrepancy between the network’s predicted output and the ground-truth annotations. This optimization was performed using Gradient Descent (GD)-based algorithms.

To identify the optimal training configuration, a k-fold cross-validation strategy was employed to select hyperparameters that are not known a priori, such as batch size, learning rate, and the number of training epochs. The training dataset was partitioned into k distinct folds. In each iteration, k-1 folds were utilized for training the network, while the remaining fold served as a validation set to monitor generalization performance. This process was repeated k times, ensuring that every fold served as the validation set exactly once. Performance metrics were computed for each iteration, and the optimal hyperparameter configuration was selected based on the highest monitored validation metrics, averaged across all k folds. Once the optimal configuration was established, the final production models were retrained on the entire training dataset to maximize training set cardinality. The technical performance of these fully trained models was subsequently assessed on an independent held-out test set to ensure unbiased evaluation.

## 4. Experimental design

### 4.1. Ethics and participants

Patients were prospectively and retrospectively enrolled between September 2019 and August 2025 in the context of a larger multicentre study, involving six clinical centers across Italy and Israel: A.O. Ordine Mauriziano di Torino (Torino, Italy), A.O.U. Città della Salute e della Scienza Ospedale Sant’Anna (Torino, Italy), Presidio Ospedaliero Ospedale Martini (Torino, Italy), IRCCS AOU Policlinico Sant’Orsola Malpighi (Bologna, Italy), Policlinico San Matteo di Pavia (Pavia, Italy), Tel Aviv Sourasky Medical Center (Tel Aviv, Israel).

The clinical trial was conducted in accordance with the Declaration of Helsinki and approved by the local institutional ethics committees of each participating center (A.O. Ordine Mauriziano di Torino, A.O.U. Città della Salute e della Scienza Ospedale Sant’Anna and Presidio Ospedaliero Ospedale Martini: Ethics Committee of A.O.U. Città della Salute e della Scienza; IRCCS AOU Policlinico Sant’Orsola Malpighi: Ethical Committee di Area Vasta Emilia Centro; Policlinico San Matteo di Pavia: Ethical Committee of Pavia; Tel Aviv Sourasky Medical Center: IRB of Tel Aviv Sourasky Medical Center).

Data collection was performed uniformly across all centers, with patients undergoing standard transvaginal and/or transabdominal US examinations according to Good Clinical Practice (GCP) guidelines. All participants signed an informed consent approved by the IRB for the use of anonymized clinical data before taking part in the study.

Eligibility criteria allowed the inclusion of women presenting (i) with no adnexal masses on gynecological examination, (ii) with a functional cyst referred for follow-up or (iii) with an adnexal mass referred for surgery or for follow-up if the lesion was stable on two consecutive assessments within 365 days and associated with negative CA-125. For adnexal masses, surgery was required within 180 days of the US examination. Patients of any age could be included, with legal guardian consent for those under 18 years; eligibility included women who underwent transvaginal or transabdominal US. Exclusion criteria were (i) absence of histological report or absence of three assessments within 365 days with negative CA-125 for non-functional masses, (ii) refusal to provide informed consent.

For the present study, a subset of 1081 adult women (Table 2) was selected from the larger multicentre cohort, according to the pre-specified inclusion and exclusion criteria. More restrictive criteria were applied to construct a dataset appropriate for the purpose of our research and compliant with the intended use of the device under validation. In particular, the pathology distribution in the selected cohort was in line with epidemiological findings, and the dataset distribution was coherent with comparable and larger international studies [80, 18], both in terms of histological distribution and ratio of malignant to benign cases. Data were acquired using different commercial sonographs (e.g. Philips, General Electrics, Samsung) to make OvAi Focus usable on different machinery.

**Table 2:** Patients characteristics for each participating center. A.O. Ordine Mauriziano di Torino (Center 1), A.O.U. Città della Salute e della Scienza Ospedale Sant’Anna (Center 2), Presidio Ospedaliero Ospedale Martini (Center 3), IRCCS AOU Policlinico Sant’Orsola Malpighi (Center 4), Policlinico San Matteo di Pavia (Center 5), Tel Aviv Sourasky Medical Center (Center 6)

| Clinical<br>center | Patients<br>number | Age | Missing<br>age | Menopause |  |  | Missing<br>menopause |
| --- | --- | --- | --- | --- | --- | --- | --- |
|  |  |  |  | Pre- | Peri- | Post- |  |
| Total | 1081 | $52.45 \pm 16.40$ | 241 | 62 | 339 | 451 | 229 |
| Center 1 | 426 | $53.78 \pm 15.71$ | 211 | 6 | 81 | 139 | 200 |
| Center 2 | 348 | $56.68 \pm 14.98$ | 11 | 41 | 105 | 197 | 5 |
| Center 3 | 5 | $31.50 \pm 14.80$ | 1 | 0 | 3 | 2 | 0 |
| Center 4 | 88 | $43.73 \pm 14.93$ | 18 | 5 | 45 | 20 | 18 |
| Center 5 | 64 | $49.95 \pm 15.81$ | 0 | 10 | 25 | 29 | 0 |
| Center 6 | 150 | $46.71 \pm 17.75$ | 0 | 0 | 80 | 64 | 6 |

### 4.2. Dataset

The full database used for algorithm training, validation and testing accounted for a total of 1081 clinical cases, including 751 cases with ovarian masses only, 182 cases with functional ovaries and 148 cases with both structures (Table 3). For each clinical case, one or more items, i.e. US videos and/or images, acquired using transvaginal or transabdominal probes were available. Clinical cases were classified according to the anatomical structures visible in their associated items. For example, cases classified as ovarian masses only included exclusively items in which an adnexal mass was visible. Histological report was associated with the large majority of cases presenting an adnexal mass (99.7 %). Clinical cases with an adnexal mass but lacking a histological report were excluded from the test set, but included in the training set.

**Table 3:** Dataset description at clinical case and item level.

| Granularity | Feature | Full dataset | Segmentation |  | Morphology |  |
| --- | --- | --- | --- | --- | --- | --- |
|  |  |  | Train | Test | Train | Test |
| Clinical case | Total number | 1081 | 745 | 259 | 554 | 200 |
|  | Ovaries only | 182 | 135 | 47 | - | - |
|  | Masses only | 751 | 499 | 175 | 536 | 178 |
|  | Both | 148 | 111 | 37 | 18 | 22 |
| Item | Total number | 2734 | 1801 | 676 | 1356 | 515 |
|  | Video | 1423 | 804 | 364 | 575 | 236 |
|  | Image | 1311 | 997 | 312 | 781 | 279 |
|  | B-mode | 1793 | 1180 | 420 | 842 | 313 |
|  | Doppler | 941 | 621 | 256 | 514 | 202 |

The dataset used for the segmentation algorithm included videos and images belonging to a total of 1004 clinical cases, splitted in training and test set (Table 3). The training set was composed of a total of 745 clinical cases, of which 499 with ovarian lesions only, 135 with functional ovaries only and 111 with both adnexal mass and functional ovary, for a total of 1801 items. The test set was composed of a total of 259 independent clinical cases, never used for training or validation purposes. Among them, 175 cases included adnexal lesions only, 47 only functional ovaries and 37 both structures, counting a total of 676 items.

The dataset used for the morphology algorithm was composed of a total of 754 clinical cases including an ovarian lesion, all associated with a histological report and US videos and/or images (Table 3). The training set contains most of the clinical cases (86 %) of the training dataset already used for the segmentation module, thus keeping the same population distribution. It counted 554 clinical cases, for a total of 1356 items. The test set consisted of 200 clinical cases, never included in training or validation datasets, associated with 515 items overall. This selection was performed to guarantee a complete non-overlap with the segmentation module’s training set, thus preserving the independence of the validation procedures. This methodological constraint prevents data leakage that could otherwise produce inflated or overly optimistic performance estimates for the segmentation module on the cases later assessed by the morphology module.

### 4.3. Evaluation procedures

The framework used to evaluate both the segmentation and morphology modules was based on a performance assessment carried out on their independent test set. As standard practice in machine learning, this allows to determine the generalization performance of the algorithm. The evaluation protocol was defined separately for each task, according to the specific output characteristics, as described in the following subsections.

The evaluation of segmentation module output was performed through a comparison with manual segmentation, which is currently the most used gold standard [81]. A clinical team was initially instructed by expert gynecologists to recognize typical anatomical features of functional ovaries and ovarian lesions according to their histological type. Then, it was trained to manually label ground-truth masks of functional ovaries, solid components of adnexal masses, and cystic components of adnexal masses using the Labelbox platform (Labelbox Inc., USA).

For each clinical case, the team examined all imaging data and metadata available - including US videos and images, US report and histological report - to determine the content of each image or video in terms of visible organs and to select the items relevant for the segmentation task. Graphical masks were applied to the selected items, while remaining areas were labelled as background. Each mask was reviewed by an expert medical doctor to guarantee the correctness of ground-truth labels. Predicted masks obtained from the segmentation algorithm were compared with ground-truth masks through the computation of standard case-level performance metrics, then aggregated across the dataset.

The validation of morphology module output was based on an inter-observer agreement analysis. In the morphological characterization of ovarian lesions based exclusively on US imaging, the most reliable reference is usually represented by the assessment performed by an expert clinician with several years of experience and extensive knowledge of IOTA terms and definitions. However, several studies have demonstrated a non-negligible variability in the interpretation of adnexal masses features even among highly experienced observers [21, 22, 23, 24], indicating that the assessment of a single observer cannot be regarded as a valid gold standard. For these reasons, inter-observer agreement was deemed more appropriate and robust for validation than traditional performance metrics such as accuracy, precision, and recall, as it allows the evaluation of consistency across observers.

The computed agreement values were compared with those reported by Meys et al. [25], whose work represents the most similar experimental setting and provides a comparable benchmark for our analysis. The test set was systematically partitioned into four distinct groups (groups A, B, C and D; n = 50 cases per group). A stratified random sampling procedure was employed to ensure a homogeneous distribution of histological types across the four groups. A panel of seventeen gynecologists from multiple contributing centers in Italy and Spain (A.O. Ordine Mauriziano di Torino, A.O.U. Città della Salute e della Scienza Ospedale Sant’Anna, Presidio Ospedaliero Ospedale Martini, Policlinico San Matteo di Pavia, Hospital Universitario Puerta de Hierro Majadahonda de Madrid), with heterogeneous levels of experience, participated in the validation as observers. Observers were assigned to the four groups in a balanced manner to ensure that each group included clinicians with different experience levels and from different centers. Each group was evaluated by a comparable number of observers (group A: n = 4, group B: n = 5, group C: n = 4, group D: n = 4).

Each observer independently evaluated one of the four groups of clinical cases. Specifically, for each clinical case, the observer had access to the relative US images and videos. After a comprehensive inspection, the observer was asked to fill a structured form providing a subjective morphological assessment with the four IOTA-based keyword ratings, compliant with Morphology output (Acoustic shadow, Margins, Lesion Type, Classification). Morphology module output was compared against the morphological descriptions provided by the observer’s panel, according to a leave-one-observer-out scheme illustrated in the Data analysis section.

### 4.4. Data analysis

According to the definition of a separate protocol for the segmentation and morphology module, also the data analysis was performed differently. This section describes the metrics and the aggregation strategies applied for the two tasks.

#### 4.4.1. Segmentation metrics

As stated in the Evaluation Procedures section, the generalization ability of the segmentation algorithm was assessed on the test set, comparing predicted masks with ground truth masks through the estimation of the following metrics:

- IoU is a standard metric used to evaluate the accuracy of an object detector or segmentation model by quantifying the overlap between two areas. Its value ranges from 0 to 1, where 0 indicates no overlap and 1 represents a perfect match between prediction and ground truth. In this case, given *A_p_* the number of pixels of the predicted mask and *A_g_* the number of pixels of the ground truth mask, the IoU was defined as:

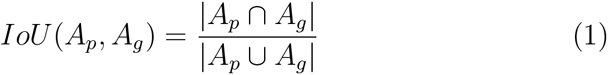

where *A_p_ ∩ A_g_* is the intersection between the two areas and *A_p_ ∪ A_g_* is the union between the two areas.

- DICE, also known as Sørensen–Dice index, quantifies the spatial overlap between two areas by measuring their similarity. The DICE score ranges from 0 to 1, where 0 indicates no overlap and 1 denotes perfect agreement. Mathematically, the DICE between predicted number of pixels *A_p_* and ground truth number of pixels *A_g_* was defined as:

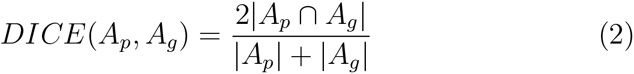

Due to its sensitivity to both false positives and false negatives, this metric is especially valuable in medical imaging, where accurate boundary delineation is crucial.

- Precision, or positive predictive value, quantifies the proportion of correctly predicted positive pixels relative to all pixels predicted as positive. This metric is defined as:

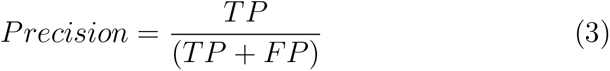

where *TP* is the number of true positives (correctly predicted foreground pixels), while *FP* is the number of false positives (background pixels incorrectly predicted as foreground).

- Recall, or sensitivity, measures the proportion of correctly predicted positive pixels relative to all actual positive pixels in the ground truth:

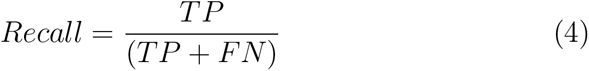

where *FN* is the number of false negatives (foreground pixels incorrectly predicted as background).

Precision and recall are typically used to assess the accuracy of predicted labels against ground truth labels. Together, they provide complementary information, as precision reflects the reliability of positive predictions, whereas recall describes the completeness of the predicted segmentation. The optimization of both metrics is relevant in segmentation tasks to ensure accurate and exhaustive outcomes.

As discussed by Reinke et al. [82], the choice of aggregation strategy for evaluating segmentation tasks has a strong impact on reported performance. According to Muller and colleagues [83], micro-aggregation computes metrics globally by pooling all pixels across the dataset, thereby weighting contributions in proportion to structure size or class frequency. Conversely, macro-aggregation computes metrics independently for each image or class and then averages them, assigning equal weight to each unit regardless of its size or prevalence. As a consequence, micro-aggregation reflects the expected performance over all pixels in the dataset, whereas macro-aggregation emphasizes the “per-case experience,” which may over-represent rare or atypical cases. In this study, we therefore adopted micro-aggregation to summarize performance across the full dataset, giving greater influence to larger, clinically relevant structures.

#### 4.4.2. Morphology agreement analysis

As described in the algorithm section, the morphology output was computed at item-level. Conversely, observers provided a single morphological description for each clinical case. In light of this, for each clinical case *c* and each IOTA-based keyword *K_j_* (with *j* = 1*, . . . ,* 4), the case-level morphology output (*CLMO*) was computed as the statistical mode of all item-level outputs associated with that case:

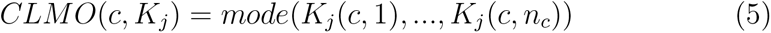

where *n_c_*is the number of items belonging to case *c*.

To quantitatively assess the agreement between *CLMO* and the observers evaluation, a specific methodological approach based on a leave-one-observer-out logic was implemented (Figure 4), following the listed steps:

1. Rating Matrices. Considering the four groups of clinical cases, indexed as *i* = 1*, . . . ,* 4 and the four IOTA-based keywords, indexed as *j* = 1*, . . . ,* 4, a Rating Matrix *RM_ij_*was constructed for each pair (*i, j*). Each matrix includes:

- the ratings for IOTA-based keyword *K_j_*on all cases of the i-th group, provided by all the *N_i_* human observers associated with that group, indexed as *h* = 1*, . . . , N_i_*. Each set of human observer ratings (*HOR*) is defined as:

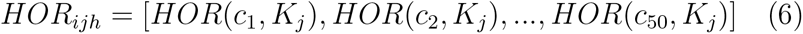
- the corresponding *CLMO_ij_*, treated as the rating of an additional observer (namely, OvAi Focus), and defined as:

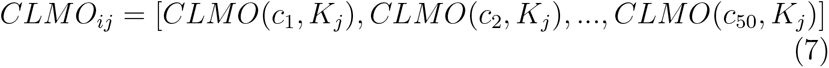 If group i has *N_i_* observers, then *RM_ij_* has (*N_i_* + 1) rows - one for each observer - and 50 columns, as the number of cases in group *i*. The process to construct each *RM_ij_* is illustrated in Figure 4.
2. Leave-One-Observer-Out (*LOO*) matrices. At this stage, each observer’s rating was removed from *RM_ij_* in an iterative, leave-one-out fashion, thus producing a set of reduced matrices 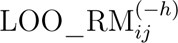. The superscript (*−h*) indicates the removal of the h-th observer from the matrix. Two types of removals were applied:

- Removal of *CLMO_ij_* (OvAi Focus): performed once for each *RM_ij_*, yielding to the matrix 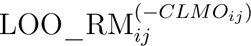, which includes only human observer’s ratings.
- Removal of a human observer: for each human observer (*h* = 1*, …, N* ) in the group *i*, a matrix 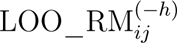 was obtained, containing all observers except the removed h-th human observer, plus OvAi Focus.
3. Agreement metrics. For each *LOO* matrix, the inter-observer agreement was assessed using both Fleiss’ Kappa coefficient (*κ*) and Percentage Agreement (*PA*), obtaining the following outputs:

- Baseline agreement, without OvAi Focus (Without-Focus). For each pair (*i, j*), 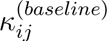 and 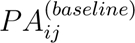 were computed from 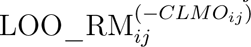, representing agreement among human observers only.
- Agreement with OvAi Focus (With-Focus-as-Observer). For each pair (*i, j*), we computed 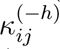 and 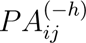. The values were obtained from 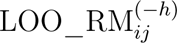.

**Figure 4:**
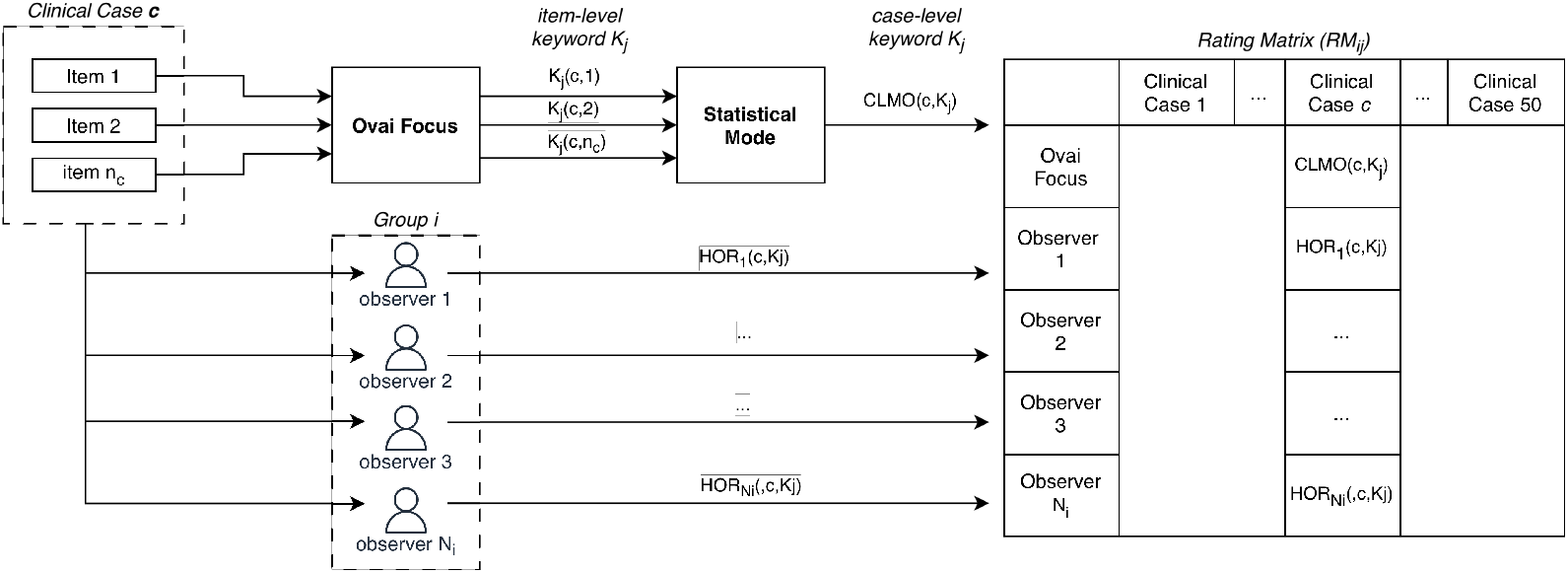
Evaluation procedure for agreement analysis

Kappa values were interpreted according to the Landis and Koch guidelines: values of 0.81-1 indicate very good agreement, 0.61–0.80 good agreement, 0.41-0.60 moderate agreement, 0.21-0.40 fair agreement, and < 0.20 poor agreement [84]. Both *κ* coefficient and *PA* were reported to allow an appropriate reading of the results and to verify the presence of the so-called Kappa Paradox phenomenon [85]. Indeed, the *κ* coefficient can be influenced by trait prevalence and marginal homogeneity, potentially leading to discrepancies when compared with *PA*. In such cases, interpretation of agreement should consider both metrics.

## 5. Results

Results obtained on the test set for the Segmentation module are reported in Table 4. Specifically, it presents the segmentation performance for the internal anatomical components of ovarian lesions (i.e., cystic and solid regions), as well as for functional ovary pixels and the complete lesion area, obtained by merging the cystic and solid regions. Performance is quantified using IoU, DICE, Precision and Recall as detailed in Data analysis section (4.4.1). The proposed method achieved DICE scores of 0.68 and 0.85 for the solid and cystic components of adnexal masses, respectively, while a DICE of 0.87 was obtained for the complete lesion. Lower performance was observed for the functional ovary, with a DICE of 0.62 and an IoU of 0.45. It can be noticed that all evaluation metrics exhibit a consistent trend across categories, indicating that the relative segmentation performance of the different regions is preserved regardless of the metric considered.

**Table 4:**
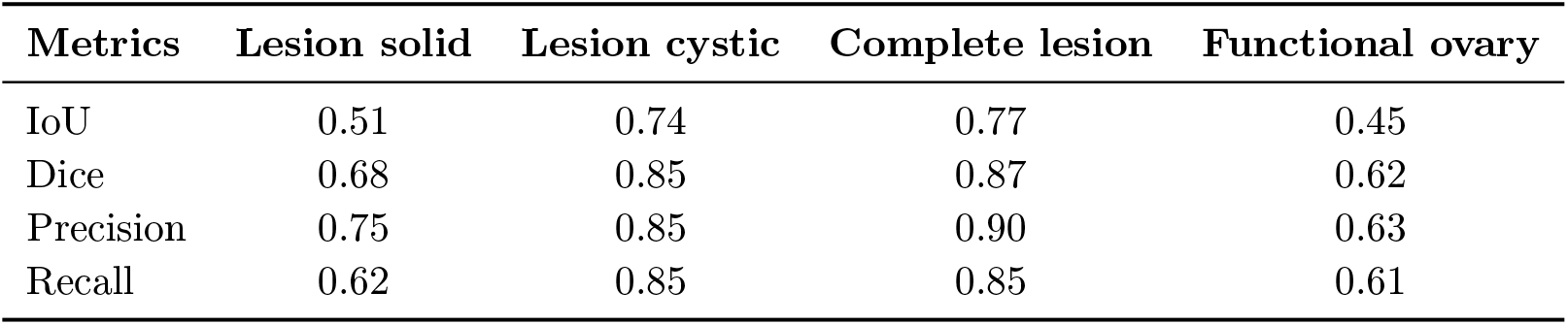
Segmentation performance metrics for different lesion components and functional ovary.

The comparison between results obtained for the Morphology module and those reported in the literature is presented in Table 5. For each IOTA-based keyword, the agreement under the Without-Focus and With-Focus-as-Observer conditions is reported in terms of *κ* and *PA*. Values are expressed as ranges (minimum-maximum across the four groups of clinical cases). In addition, literature values and corresponding 95 % confidence intervals from the study by Meys et al. [25] are provided as reference for both metrics for all IOTA-based keywords, with the exception of Lesion Type, for which no literature data in a comparable setting were available.

**Table 5:** Results of agreement analysis on the full test set in terms of Fleiss’ Kappa (*κ*) and Percentage Agreement (*PA*), with and without Focus. The absence of literature reference values for Lesion Type is indicated as N/A.

| IOTA-based<br>keywords | Literature ranges |  | Without-Focus |  | With-Focus |  |
| --- | --- | --- | --- | --- | --- | --- |
| | $\kappa$ (95% CI) | $PA$ | $\kappa$ | $PA$ | $\kappa$ | $PA$ |
| Acoustic Shadows | 0.34 (0.06–0.62) | 74 % | 0.33–0.46 | 84–90 % | 0.26–0.29 | 80–86 % |
| Margins | 0.41 (0.18–0.63) | 66 % | 0.14–0.47 | 61–75 % | 0.16–0.42 | 60–72 % |
| Classification | 0.39 (0.24–0.53) | 52 % | 0.36–0.51 | 50–61 % | 0.32–0.38 | 47–51 % |
| Lesion Type | N/A | N/A | 0.34–0.67 | 56–78 % | 0.35–0.57 | 57–72 % |

The same results are graphically presented in Figure 5 and 6. Figure 5 shows the inter-observer agreement in terms of *κ* for each clinical case group (*i* = 1*, . . . ,* 4) and each IOTA-based keyword (*j* = 1*, . . . ,* 4). Each sub-plot highlights the difference between the Without-Focus and With-Focus-as-Observer conditions. Literature values from Meys et al. [25], together with their 95 % confidence intervals, are outlined as dashed lines and shaded areas, when available. Figure 6 presents the same information as Figure 5, expressed in terms of *PA*, with literature reference values indicated by dashed lines, when available.

**Figure 5:**
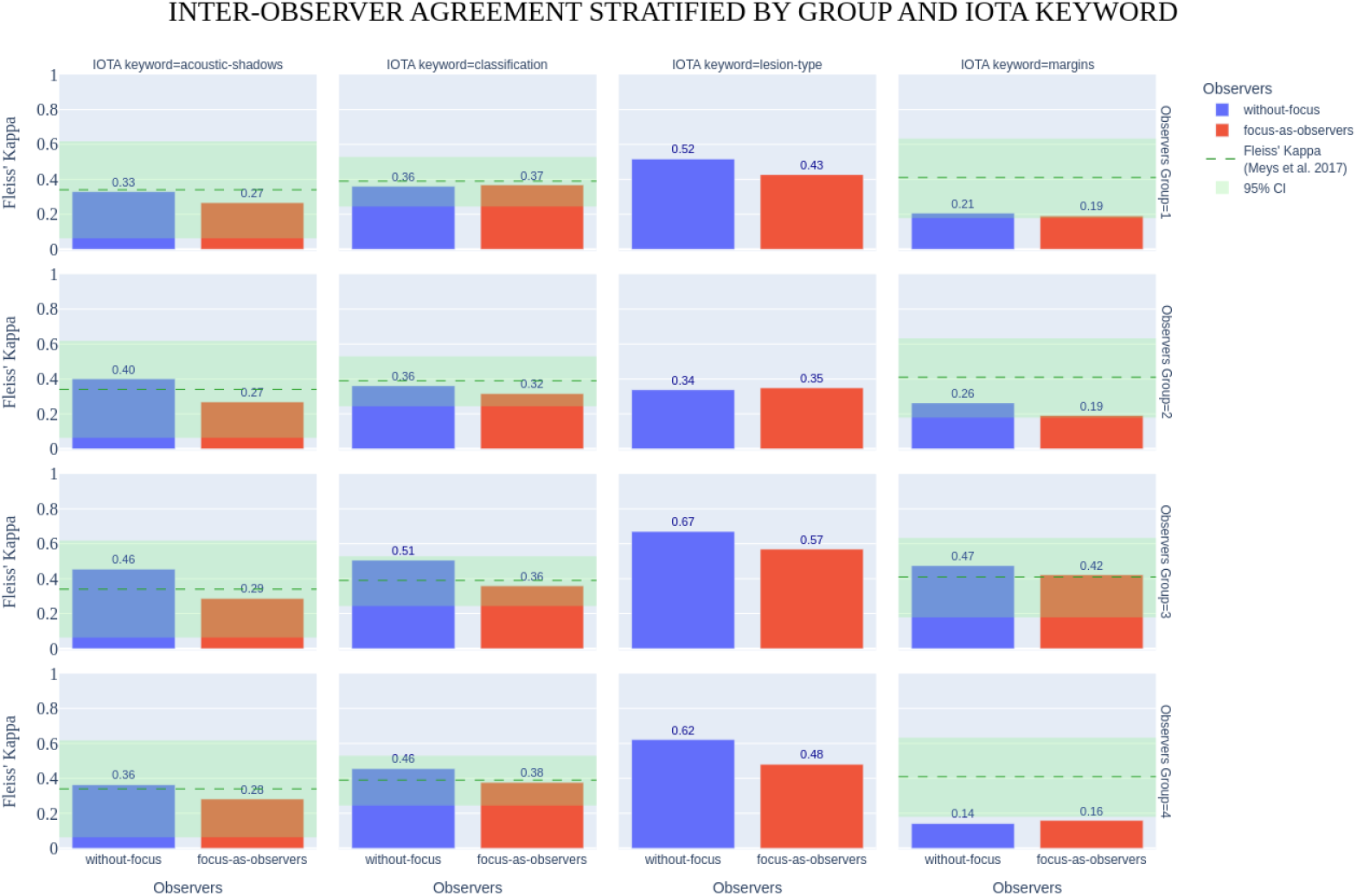
Results of Mophology module on the full Test Set in terms of Fleiss’ Kappa coefficient for each group and IOTA-based keyword: Without-Focus (blue), Focus-as-Observer (red), Meys et al. reference value and 95 % confidence interval (green dashed line and shaded areas, respectively).

**Figure 6:**
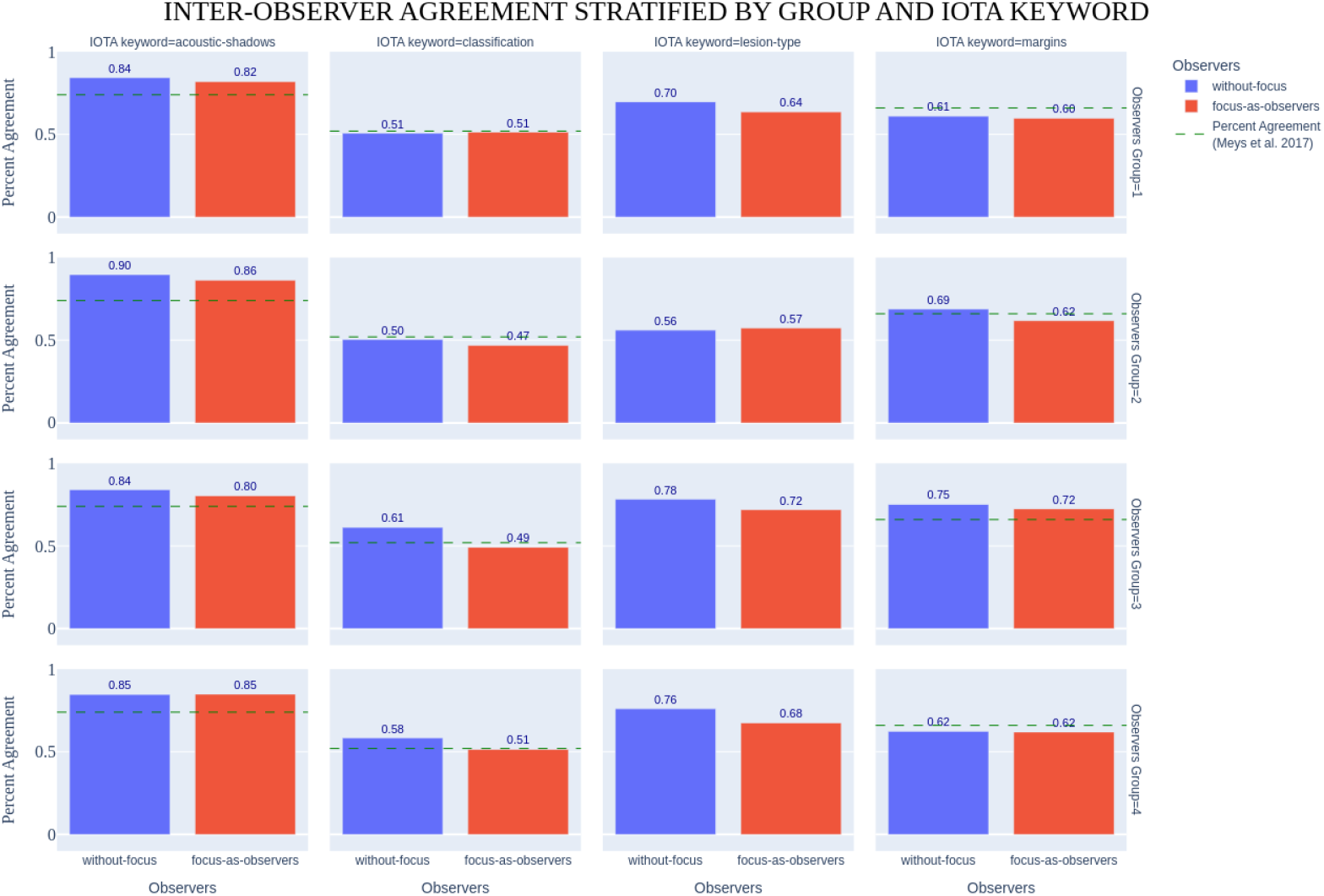
Results of Mophology module on the full Test Set in terms of Percentage Agreement for each group and IOTA-based keyword: Without-Focus (blue), Focus-as-Observer (red), Meys et al. reference value (green dashed line).

Across the four clinical case groups, the inter-observer agreement showed variability depending on the IOTA-based keyword. Under the Without-Focus condition, *κ* values ranged from 0.33 to 0.46 for Acoustic Shadows, from 0.14 to 0.47 for Margins, from 0.36 to 0.51 for Classification, and from 0.34 to 0.67 for Lesion Type. Regarding With-Focus-as-Observer condition, the corresponding *κ* ranges were 0.26 - 0.29 for Acoustic Shadows, 0.16 - 0.42 for Margins, 0.32 - 0.38 for Classification, and 0.35 - 0.57 for Lesion Type.

*PA* values followed a similar pattern. In the Without-Focus condition, PA ranged from 84 % to 90 % for Acoustic Shadows, from 61 % to 75 % for Margins, from 50 % to 61 % for Classification, and from 56 % to 78 % for Lesion Type. In the With-Focus-as-Observer condition, PA values ranged from 80 % to 86 % for Acoustic Shadows, 60 % to 72 % for Margins, 47 % to 51 % for Classification, 57 % to 72 % for Lesion Type.

Overall, when considering *κ*, Lesion Type achieved the highest values across conditions. In contrast, Acoustic Shadows displayed the highest raw agreement in terms of PA. Across all IOTA-based keywords, *κ* values were generally lower and more variable than *PA*.

## 6. Discussion

The metrics shown for the Segmentation module (Table 4) highlight an overall good performance, in line with or superior to those reported for comparable state-of-the-art solutions, confirming the robustness and accuracy of the implemented algorithms.

The method achieved its best results in segmenting the cystic components of adnexal masses, reaching a DICE of 0.85 and an IoU of 0.74. This out-come can be attributed to the US characteristics of cystic regions, which are typically characterized by more homogeneous content and clearer boundaries with respect to surrounding tissues, thus facilitating their correct detection. Conversely, the segmentation of solid components appeared to be more challenging, with a Dice score of 0.68 and an IoU of 0.51. Compared to cystic regions, solid components show more heterogeneous echogenic patterns and less well-defined borders, and can often be confused with surrounding structures, affecting segmentation accuracy. To the best of our knowledge, reference values for the segmentation of solid and cystic components of adnexal masses are currently not available in the literature, highlighting the novelty of the proposed approach.

The complete lesion area, obtained by merging solid and cystic components, showed the highest performance (Dice score of 0.87, IoU of 0.77), proving that the model is able to capture the overall size of the adnexal mass in a reliable way, which is of primary importance for diagnosis and treatment planning [86]. For this task, results reported in the literature were in line with our findings: Dai-Li et al. [37] reported an average Dice of 0.89 on an internal test set and 0.82 on an external test set, while comparable performance was reported by Barcroft et al. [36] (average Dice 0.85) and Whitney et al. [45] (median Dice 0.91).

Lower performance was observed for functional ovary segmentation, with a Dice score of 0.62 and an IoU of 0.45. This result is reasonable considering the characteristics of our dataset, which includes a higher prevalence of adnexal masses compared to functional ovaries (30 % cases with functional ovaries versus 83 % with adnexal masses). Moreover, from the clinical perspective, the lack of sharp anatomical boundaries between the functional ovary and the surrounding tissues, together with the high inter-patient variability in ovarian morphology, may negatively affect segmentation performance for this category. Despite this, the Dice score obtained by OvAi Focus on both images and videos fall within the uncertainty range reported by Dai-Li and colleagues on static images (DICE of 0.69 ± 0.26 on an internal test set and 0.68 ± 0.27 on an external test set).

Overall, all evaluation metrics exhibited a consistent trend across categories, indicating that the relative segmentation performance is independent of the metric adopted. This behaviour supports the robustness of the proposed solution, suggesting that the observed differences are mainly related to the US characteristics of the segmented structures.

Based on current information, no studies in the literature describe automated methods for IOTA-based keywords extraction directly comparable to our approach. In current clinical practice, subjective inter-observer agreement among human readers is considered the gold standard for the morphological description of adnexal masses according to IOTA definitions. Several studies have assessed such agreement using Cohen’s or *κ* coefficients [25, 26, 27, 21, 22, 23, 24], reporting a high variability in agreement across keywords and observers’ experience.

In this context, the study of Meys and colleagues [25] was selected as a benchmark for comparison, being the closest experimental setting to the present work. Literature agreement values from Meys et al. were therefore reported alongside the results of this study, summarized as mean values and 95 % CIs in Table 9 and graphically reported in Figures 9 and 10.

The results of the present study confirm a marked heterogeneity in agreement levels across IOTA-based keywords and clinical case groups, in line with existing literature [25, 26, 27, 21, 22, 23, 24]. Considering the Without-Focus condition, *κ* values ranged from 0.14 to 0.67 across the evaluated keywords, showing high convergence with the ranges from Meys et al. [25]. Under With-Focus-as-Observer condition, agreement values slightly decreased, with *κ* values ranging from 0.16 to 0.57; however, these values remained within the reported 95 % CIs, indicating consistency with expected human inter-observer variability. A comparison between *κ* and *PA* revealed a non-negligible discrepancy between the two metrics. In particular, *PA* values were consistently higher than *κ*, ranging from 47 % to 90 % depending on the condition (Table 5). This behavior is consistent with the “Kappa paradox” described in Section 4.4.2 and supports the joint interpretation of both metrics. Across all conditions, Lesion Type showed the highest interobserver agreement in terms of Fleiss’ *κ*, reaching values up to 0.57 under the With-Focus-as-Observer condition. In contrast, Acoustic Shadows achieved the highest Percentage Agreement, with PA values consistently above 80 % and reaching up to 90 % in the Without-Focus condition. Classification exhibited comparatively lower agreement, particularly under the With-Focus-as-Observer condition (*κ* 0.32-0.38; PA 47-51%). This finding is expected, as Classification involves a larger number of categories compared with other IOTA-based keywords, inherently reducing the likelihood of consensus. Margins demonstrated moderate agreement (*κ* 0.16-0.42 under With-Focus-as-Observer), while Lesion Type showed greater variability across conditions.

In general, although the inclusion of OvAi Focus as an observer resulted in slightly lower agreement compared to human-only panels, the observed values remain comparable to literature reference standards. These findings indicate that the objective assessment provided by the Morphology module of OvAi Focus aligns with the expected inter-observer variability among expert readers, supporting its potential role as a reliable tool for IOTA-based morphological evaluation.

## 7. Conclusion

OvAi Focus is a novel AI-based medical software designed for multi-class segmentation of adnexal structures and IOTA-compliant morphological characterization in gynecological US. This study demonstrates its unique ability to simultaneously delineate functional ovaries and adnexal masses, distinguishing cystic and solid components, while automatically extracting clinically relevant morphological descriptors. Segmentation performance was robust on an independent multicenter test set, particularly for the whole lesion area and cystic components, aligning with state-of-the-art literature. Although segmentation performance for functional ovaries and solid components was lower, OvAi Focus is the first validated framework capable of this dual task, representing a major advance in AI-assisted gynecological US. Morphology extraction was validated through inter-observer agreement with seventeen gynecologists, showing performance comparable to trained human readers and potential to reduce subjectivity in adnexal mass assessment. Based on our knowledge, this is the first automatic approach proposed for the characterization of adnexal masses. Strengths include multicenter design, independent validation sets, strict prevention of data leakage, and alignment with internationally recognized IOTA terminology. Limitations involve geographic dataset scope and morphology validation based on agreement rather than histopathology. Future work will focus on expanding dataset diversity, integrating the software into prospective clinical workflows, and extending automated extraction to additional IOTA-based features. OvAi Focus represents a step toward standardized, objective, and reproducible US characterization of adnexal structures, supporting clinicians and improving consistency in gynecological assessment.

## Data Availability

The data that support the findings of this study are the property of SynDiag s.r.l. and are not publicly available. The results generated from these data are reported in this manuscript.

## 8. Declaration of Interest

RBM, DC and FG hold stock in SynDiag, where they also have paid leadership roles. FS, NT and PRF are employees of SynDiag. The device evaluated in this study is a CE-marked medical device developed and commercialized by SynDiag. SynDiag had a role in the study design, data analysis, and manuscript preparation. The remaining authors declare no competing interests.

## 9. Declaration of generative AI use

The authors used ChatGPT exclusively to check grammar and typos.

## Notes

### Author Declarations

Ethics Committee of A.O.U. Citta della Salute e della Scienza gave ethical approval for this work Ethical Committee di Area Vasta Emilia Centro gave ethical approval for this work Ethical Committee of Pavia gave ethical approval for this work IRB of Tel Aviv Sourasky Medical Center gave ethical approval for this work

